# Right Anterior Temporal Degeneration, Emotions, and Loss of Nonverbal Semantics: The Emotional Semantic Variant Frontotemporal Dementia

**DOI:** 10.1101/2021.06.07.21258432

**Authors:** Kyan Younes, Valentina Borghesani, Maxime Montembeault, Salvatore Spina, Maria Luisa Mandelli, Ariane E. Welch, Elizabeth Weis, Patrick Callahan, Fanny M. Elahi, Alice Y. Hua, David C. Perry, Anna Karydas, Daniel Geschwind, Eric Huang, Lea T. Grinberg, Joel H. Kramer, Adam L. Boxer, Gil D. Rabinovici, Howard J. Rosen, William W. Seeley, Zachary A. Miller, Bruce L. Miller, Virginia E. Sturm, Katherine P. Rankin, Maria Luisa Gorno-Tempini

## Abstract

It has been proposed that focal anterior temporal lobe (ATL) degeneration is a specific, unitary FTLD-TDP-related disease that initially preferentially affects the left or right hemisphere. Patients with early left ATL (lATL) atrophy show severe anomia and verbal semantic deficits and meet criteria for semantic variant primary progressive aphasia (svPPA) and semantic dementia, prompting appropriate neurological care. There is less consensus regarding the symptoms in right ATL (rATL) predominant cases, who most often present with behavioral and emotional changes leading to a misdiagnosis of a psychiatric disorder, and later of behavioral variant frontotemporal dementia (bvFTD). Uncertainties regarding early symptoms and lack of an overarching framework continues to hinder proper diagnosis and care of patients with rATL disease. Here, we present symptom chronology, cognitive and socioemotional profiles of a large, well-characterized, longitudinally followed cohort of patients with rATL-predominant degeneration and propose new criteria and nosology for the syndrome.

We identified individuals with a clinical diagnosis of bvFTD or svPPA and a structural MRI (n=478). Based on neuroimaging criteria, we identified three groups: patients with rATL-predominant atrophy with relative sparing of the frontal lobes (n=46), patients with frontal-predominant atrophy with relative sparing of the rATL (n=79), and patients with lATL-predominant atrophy with relative sparing of the frontal lobes (n=75). Seventy-eight patients had undergone autopsy. We analyzed patients’ clinical, neuropsychological, genetic, anatomical, and pathological profiles.

In the rATL-predominant group, the earliest symptoms were loss of empathy (27%), person-specific semantic impairment (23%), and complex compulsions and rigid thought process (18%). On testing, this group exhibited greater impairments in emotional theory of mind, identifying famous people from names and face, and facial affect naming (despite preserved face perception) than the lATL- and frontal-predominant groups. The clinical features were highly sensitive (81%) and specific (84%) in differentiating rATL from bvFTD in the first three years of the disease. FTLD-TDP (84%) was the most common pathology.

Our results suggest that rATL-predominant degeneration is characterized by early loss of empathy and person-specific knowledge, deficits that are caused by progressive loss of semantic memory for concepts of social-emotional relevance. Although this syndrome exists along a clinical, anatomical, and pathological continuum with svPPA, patients present with progressive behavioral changes. To facilitate early identification and care in clinical and research settings, we propose specific diagnostic criteria and the term “emotional semantic variant frontotemporal dementia” to distinguish this syndrome from other forms of frontotemporal dementia.

## Introduction

The term “frontotemporal dementia” (FTD) was introduced to encapsulate the progressive personality changes, social conduct impairment, and language deficits associated with atrophy of the frontal and temporal lobes.^1^ Within FTD, behavioral symptoms often localize to frontal, temporal, insular, and striatopallidal regions in the right hemisphere, whereas language deficits typically localize to structures in the left.^2^ Currently, the behavioral syndrome associated with FTD is referred to as “behavioral variant frontotemporal dementia” (bvFTD), and the language syndromes are brought together under the term “primary progressive aphasia” (PPA).

The syndromes that target the anterior temporal lobe (ATL) are often initially asymmetric, with focal atrophy targeting either the right ATL (rATL) or left ATL (lATL),^4,5^ and thus have distinct early clinical presentations. Patients with lATL-predominant atrophy typically have notable language deficits, which were well-represented in the consensus clinical criteria for semantic dementia and more recently for semantic variant PPA (svPPA).^6,7^ Although these criteria do include semantic deficits for objects and faces, they primarily emphasize verbal semantic deficits that result in anomia, single word comprehension deficits, and object-identification impairments, and do not highlight socioemotional and behavioral deficits and thus overlooking the important symptoms resulting from rATL degeneration.

Patients with focal atrophy of the rATL, exhibit prominent emotional changes and behavioral symptoms that can be hard to distinguish from those of bvFTD, and may not initially have significant aphasia symptoms.^4,8–13^ Although in the early stages of disease, atrophy may be asymmetric and target the ATL unilaterally, over time the disease spreads to the contralateral hemisphere, and language and behavioral symptoms converge.^4,13,14^ As both lATL- and rATL-predominant degeneration are typically associated with frontotemporal lobar degeneration - transactive response DNA binding protein 43 type C (FTLD-TDP type C) pathology,^5^ lateralized ATL presentations are thought to reflect different manifestations of a single pathological continuum.^4,15–19^ Studies that have investigated the clinical characteristics of rATL-predominant degeneration find that patients show difficulties recognizing familiar people and empathizing with others.^4,11,20–22^ Diminished empathy—which can include lack of emotional responsiveness as well as decreased social connection and compassion—is often remarkable when disease targets the rATL.^11,23–26^ Neuroimaging studies have associated rATL atrophy with deficits in a wide range of socioemotional functions including empathy,^23^ nonverbal social cue (e.g., sarcasm) detection,^27^ and facial emotion recognition.^28,29^ Feelings of familiarity that known others typically elicit may also rely preferentially on the rATL,^28^ which suggests that even friends and family members may lose their affective significance to patients with rATL-predominant dysfunction.

By integrating information from primary and association sensory and motor cortices, the ATLs are considered amodal hubs that represent all categories of semantic knowledge.^15,18,30–34^ Though there is strong evidence for bilateral ATL contribution to semantic conceptual knowledge, specialization is hypothesized to appear as a result of divergent inputs from right and left hemispheres.^19,35–38^ Whereas the lATL would bind verbal features into semantic conceptual knowledge through strong connections with linguistic networks, the rATL may be more centrally involved in representing nonverbal semantic knowledge through its prominent connections with right-sided socio-emotional networks.^16,32,39,40^ According to this framework, lATL degeneration would disrupt verbal semantic knowledge, such as word comprehension and retrieval, and rATL degeneration would degrade nonverbal, socially-relevant semantic knowledge in individuals with typical hemispheric functional lateralization. Consistent with this hypothesis, nonverbal semantic knowledge tasks, including visual semantic associations;^41^ identification of living beings (animals are recognized mainly by their visual features);^39^ sound recognition;^42^ and tactile,^17^ olfactory,^43^ and gustatory stimulus recognition have all been linked to rATL.^44^ Importantly, nonverbal semantic processing is also necessary for the recognition and identification of familiar and famous people from faces and voices.^28,42^ By extension, the rATL is also hypothesized to link emotion concepts with visual associations as well as non-verbal bodily changes (e.g., changes in autonomic nervous system functioning, motor activity, and experience), thus serving as the core hub for socioemotional semantic processing.^31,45,46^ Although patients with rATL-predominant degeneration are often described as having “prosopagnosia,” this term does not fully capture the deficit, because patients cannot recognize familiar people from visual (face), linguistic (name), or auditory cues (voices), indicating a broader semantic deficit for biographical, person-specific semantic knowledge.^47^ This lateralized specialization is observed in individuals known to have left-brain language dominance, non-right-handed patients can present with reverse symptomatology if they have reversed hemispheric dominance.^25,48^

Despite these advances in theoretical understanding of right and left ATL functions, studies systematically testing both semantics and socioemotional functions in large well-characterized cohort of patients are still lacking. Furthermore, there is no consensus on a set of diagnostic criteria for the rATL-predominant syndrome, and patient symptomatology has not been clearly linked to theoretical cognitive models. Patients with rATL-predominant atrophy pose a nosologic challenge as they can have symptoms that overlap with the diagnostic criteria for svPPA and semantic dementia,^6,49^ which emphasize verbal semantic deficits, and bvFTD, which focus on behavioral and emotional features.^50^ Lacking formal nomenclature or diagnostic criteria, patients with rATL-predominant degeneration are described with terms such as right temporal svPPA, right temporal semantic dementia, right temporal bvFTD, and right temporal variant FTD.^4,8–10,47,51–53^ Because loss of empathy is often misinterpreted as a psychiatric symptom and because there are no clinical criteria for the rATL-predominant syndrome, these patients are often identified later in the disease course, when severe behavioral impairment often justifies a diagnosis of bvFTD. Furthermore, the current conflation of rATL-predominant and bvFTD syndromes may cause unnecessary confusion about patients’ underlying neuropathology, which is much more heterogeneous in bvFTD, and thus will make the selection of a disease-modifying treatment more difficult for clinicians as pathology-specific drugs become common in the near future. Diagnostic criteria that would facilitate the early identification of rATL-predominant patients would also accelerate studies of nonverbal semantics and allow for the development of reliable measures that track these socioemotional changes in neurodegenerative illnesses.

The goal of the present study was to examine the clinical, neuropsychological, genetic, anatomical, and pathological characteristics of a large cohort of patients with rATL-predominant atrophy. Patients were studied within a multidisciplinary project on FTD-spectrum disorders that included comprehensive assessments of both language and socioemotional functioning. Based on the results, we propose new diagnostic criteria for a rATL-predominant syndrome that is on a continuum with, but qualitatively and quantitatively distinct from, both bvFTD and lATL-dominant svPPA syndromes as currently defined. To evaluate the validity of these criteria, we compared these rATL patients to those with frontal-predominant bvFTD and lATL-predominant svPPA, as determined by clinical and neuroimaging criteria. We hypothesized that patients with rATL-predominant damage would have a specific pattern of early semantic memory loss for socio-emotionally relevant, non-verbal concepts, such as famous people and emotions, also resulting in typical behavioral symptoms such as loss of empathy. We expected that lack of empathy would be a prominent feature, and that other bvFTD behavioral symptoms (e.g., disinhibition, apathy/inertia, and lack of judgment/dysexecutive symptoms) would be less common. We also anticipated that patients with rATL-predominant degeneration would have some characteristic symptoms of svPPA (e.g., word comprehension and confrontational naming difficulties) but that these would be comparatively mild and would often not meet general PPA diagnostic criteria (i.e., aphasia as the most prominent early clinical feature and the principal cause of functional impairment).

## Materials and methods

### Participants

We identified patients who met bvFTD and/or svPPA criteria (see below) and had research visits between 1998 and 2019 (n = 682) at the University of California, San Francisco (UCSF) Memory and Aging Center (MAC). As symptoms were often mild at early research visits, scores on the Clinical Dementia Rating scale (CDR) were not used when determining study inclusion.^54^ Patients who did not have a brain MRI within one year of the first research evaluation were excluded (n = 204). From the remaining 478 cases, we used structural neuroimaging measures to identify individuals with predominant rATL atrophy and relative preservation of the frontal lobes (see details below) (n = 46). We also included three other groups for comparison: a group of individuals with lATL-predominant atrophy and relative preservation of the frontal lobes (n = 75), a group of individuals with frontal-predominant atrophy and relative preservation of the rATL (n = 79), and a group of healthy older controls from the MAC Hillblom Healthy Aging Network (n = 59). We used strict clinical and anatomical inclusion criteria to contrast the rATL patients with these three groups and to clarify the distinct cognitive-behavioral phenotype of the rATL-predominant syndrome. Patients or caregivers provided informed consent following procedures aligned with the Declaration of Helsinki, and the study was approved by the UCSF Committee for Human Research.

### Diagnostic criteria

Two raters, a behavioral neurologist (KY) and a neuropsychologist (MM), reviewed all available medical data for the rATL-predominant patients to determine whether they met the following diagnostic criteria: 1) Neary-FTD,^6^ 2) Neary-Semantic,^6^ 3) bvFTD,^50^ and 4) svPPA.^49^ We also noted whether patients had semantic variant features (i.e., impaired confrontation naming and single-word comprehension) regardless of meeting PPA general criteria (i.e., aphasia is the most prominent deficit in early disease).^49^ This allowed us to describe verbal semantic deficits in patients who had predominantly behavioral presentations. The two raters determined whether each of these criteria was met at three different time points: 1) within the first three years of disease onset, 2) at the first MAC research evaluation, and 3) in the years subsequent to the first MAC evaluation.

### Detailed symptom taxonomy and chronology

All research participants were evaluated by a behavioral neurologist, a neuropsychologist, a speech and language pathologist, and a nurse. A clinical history was obtained from each patient, with corroboration from the caregiver/informant, and began by identifying the nature and onset of the first symptoms. This was followed by a chronological history of how symptoms evolved, and then a detailed inventoried review of the domains of memory, language, executive function, visuospatial abilities, behavior, sleep, sensory processing, and motor function. Patients did not need to present for evaluation at the same stage of the disease in order for the retrospective interview to fully capture the chronology of symptoms.

We documented each patient’s first five symptoms, rather than all symptoms ever noted, because we expected many of the canonical bvFTD symptoms (disinhibition, apathy, loss of empathy, compulsions, hyperorality, and executive deficits) and PPA symptoms (language and semantic impairment) would emerge for most people in the disease’s later stages. In an effort to refine our categorization of the behavioral and emotional symptoms, we catalogued symptoms according to the following taxonomy:

1. Loss of empathy: Difficulty recognizing, understanding, or responding to others’ emotions and needs; selfishness; emotional distance from others; reduced or inappropriate emotional expressivity, diminished social interest, interrelatedness, or personal warmth.
2. Words and object semantic loss: Loss of knowledge about words, facts, concepts, animate or inanimate objects, places, or landmarks. Patients may demonstrate impaired naming, diminished recall, poor identification, or reduced feelings of familiarity for these domains.
3. Person-specific semantic knowledge loss: Loss of knowledge about known faces, proper names, and people (including biographical information about famous people, close friends, and/or family members). Patients may demonstrate impaired naming, diminished recall, poor recognition, or reduced feelings of familiarity for these domains.
4. Complex compulsions and rigid thought process: Adhering to fixed schedules or roles, preoccupation with dogmas (e.g., hyper-religiosity) or health (hypochondriasis), restricted preference for certain colors, clothing, or diet, spending hours playing word games and puzzles.
5. Simple repetitive behaviors, hoarding, or obsessions: repetitive motor (e.g., clicking, tapping, pacing) or verbal stereotypies, hoarding, or preoccupation with objects or people.
6. Apathy/inertia: Cognitive (reduced planning and voluntary action), behavioral (reduced self-initiated thoughts and behaviors), and affective (reduced social, emotional, behavioral interest) forms of apathy.^55^
7. Disinhibition: Impulsivity or socially inappropriate behavior, loss of manners or decorum.
8. Lack of judgment/dysexecutive: Rash or careless actions, judgment mistakes that are out of character. Of note, in the current bvFTD criteria,^49^ lack of judgment is considered as a part of disinhibition, but for this study we separated these two symptoms as they may be subserved by different neuroanatomical systems.^56^
9. Episodic memory loss: Difficulty remembering recent events and autobiographical information.
10. Hyperorality or dietary changes: Altered food preferences, binge eating, increased consumption of alcohol or cigarettes, oral exploration, or consumption of inedible objects.
11. Motor neuron disease signs: Bulbar and limb signs of motor neuron disease.
12. Other symptoms: Visuospatial difficulties, declined hygiene, loss of sexual desire, dietary changes (increased or decreased eating), weight gain, weight loss, hypersomnia, and insomnia. These symptoms are either common in other neurodegenerative illnesses or not specific for a single neurodegenerative disease.

### Functional, cognitive, and behavioral assessments

Patients underwent a comprehensive multidisciplinary assessment that included functional, neuropsychological, and socioemotional measures, Table 1 and Table 2, as previously described.^11,57^ A description of the cognitive battery and further details about patients’ performance are presented in the Supplementary material. Verbal semantic knowledge was evaluated with the Peabody Picture Vocabulary Test (PPVT; patients were asked to choose the picture that best describes a word),^58^ the abbreviated 15-item Boston Naming Test (BNT; patients were asked to name different drawings),^59^ and semantic verbal fluency (patients generated as many animals as possible in 60 seconds). Nonverbal semantic knowledge was tested with the picture version of the Pyramids and Palm Trees (PPT-P; patients matched semantically associated pictures).^60^

**Table 1:** Demographics, functional, and cognitive scores.

|  | Right Temporal | Frontal | Left Temporal | Healthy Control |
| --- | --- | --- | --- | --- |
| **Epidemiology and Functional Scales** |  |  |  |  |
| Handedness, (right, left, ambidextrous) | 39, 6, 1 | 72, 3, 3 | 68, 6, 1 | 54, 5, 0 |
| Sex, (female, male) | 23, 23 | 39, 40 | 35, 40 | 35, 24 |
| Ethnicity, (n) | 42W, 4A | 70W, 8A, 1N | 73W, 3A | 58W, 1A |
| Age of onset, mean (sd, n) | 60.2 (8.2, 46) | 56.7 (10.0, 79) | 58.7 (7.2, 75) | NA |
| Age at evaluation, mean (sd, n) | 65.11 (7.6, 46)<sup>a</sup> | 60.75 (8.8, 79)<sup>d</sup> | 63.12 (7.0, 75)<sup>f</sup> | 74.05 (9.7, 59)<sup>a,d,f</sup> |
| Years of education, mean (sd, n) | 15.78 (3.0, 46) | 15.59 (2.9, 79)<sup>d</sup> | 16.43 (2.8, 74) | 17.07 (2.3, 58)<sup>d</sup> |
| CDR score, mean (sd, n), max = 3 | 0.97 (0.5, 45)<sup>a,c</sup> | 0.98 (0.7, 79)<sup>d,e</sup> | 0.67 (0.3, 69)<sup>c,e,f</sup> | 0.0 (0.0, 59)<sup>a,d,f</sup> |
| CDR Box Score, mean (sd, n), max = 18 | 5.5 (3.5, 45)<sup>a,c</sup> | 5.73 (3.9, 79)<sup>d,e</sup> | 3.551 (2.0, 69)<sup>c,e,f</sup> | 0.01 (0.1, 59)<sup>a,d,f</sup> |
| NPI Total (severity*frequency), EMM (se, n), max = 144 | 36.0 (20.4, 45)<sup>a,b,c</sup> | 44.7 (21.0, 46)<sup>b,d,e</sup> | 21.5 (16.2, 24)<sup>c,e,f</sup> | 8.1 (8.5, 6)<sup>a,d,f</sup> |
| NPI Caregiver Distress Total, EMM (se, n), max = 60 | 14.9 (9.3, 45)<sup>a,b,c</sup> | 19.5 (8.3, 47)<sup>a,b,c</sup> | 12.5 (7.3, 22)<sup>a,b,c</sup> | 2.5 (1.7, 9)<sup>a,b,c</sup> |
| **Global Cognition** |  |  |  |  |
| MMSE, EMM (se, n), max = 30 | 26.46 (0.87, 46)<sup>b,c</sup> | 23.39 (0.70, 77)<sup>b,d</sup> | 21.81 (0.69, 71)<sup>c,f</sup> | 27.02 (0.95, 58)<sup>d,f</sup> |
| **Visuospatial Processing** |  |  |  |  |
| Benson complex figure – copy, EMM (se, n), max = 17 | 14.72 (0.33, 46) | 13.85 (0.27, 71)<sup>e</sup> | 15.60 (0.28, 70)<sup>e</sup> | 14.61 (0.27, 36) |
| VOSP Number Location, EMM (se, n), max = 10 | 8.84 (0.26, 42)<sup>b</sup> | 7.79 (0.22, 62)<sup>b,d,e</sup> | 9.36 (0.24, 56)<sup>e</sup> | 9.01 (.27, 59)<sup>d</sup> |
| **Episodic Memory** |  |  |  |  |
| CVLT 30" short delay free recall, EMM (se, n), max = 9 | 3.93 (0.29, 43)<sup>a,c</sup> | 4.70 (0.23, 65)<sup>d,e</sup> | 2.83 (0.30, 64)<sup>c,e,f</sup> | 7.30 (0.53, 13)<sup>a,d,f</sup> |
| CVLT 10' long delay free recall, EMM (se, n), max = 9 | 2.48 (0.32, 43)<sup>a</sup> | 3.55 (0.26, 65)<sup>d,e</sup> | 1.84 (0.26, 64)<sup>c,f</sup> | 7.45 (0.59, 13)<sup>a,d,f</sup> |
| CVLT Recognition, EMM (se, n), max = 9 | 6.44 (0.26, 43)<sup>a,b</sup> | 7.86 (0.21, 64)<sup>b,e</sup> | 6.31 (0.22, 62)<sup>c,f</sup> | 8.28 (0.49, 13)<sup>a,f</sup> |
| Benson complex figure – delay, EMM (se, n), max = 17 | 5.62 (0.57, 44)<sup>a,b</sup> | 8.82 (0.51, 69)<sup>b,d</sup> | 7.36 (0.48, 75)<sup>f</sup> | 11.57 (0.75, 36)<sup>a,d,f</sup> |
| **Executive Functioning** |  |  |  |  |
| Digit Span – backward, EMM (se, n) | 4.97 (0.17, 46)<sup>b,c</sup> | 3.32 (0.14, 69)<sup>b,d,e</sup> | 4.73 (1.51, 66)<sup>c,e,f</sup> | 5.36 (1.90, 55)<sup>d,f</sup> |
| Stroop (correct in 60 seconds), EMM (se, n) | 42.80 (2.95, 26)<sup>b</sup> | 29.45 (2.21, 46)<sup>b,e</sup> | 37.98 (2.12, 49)<sup>e</sup> | NA |
| Trails (Time), EMM (se, n), max = 120" | 61.40 (4.53, 42)<sup>a,c</sup> | 73.10 (3.95, 59)<sup>d,e</sup> | 45.23 (3.98, 61)<sup>c,e</sup> | 32.87 (4.60, 59)<sup>a,d</sup> |
| Design fluency, EMM (se, n) | 6.50 (0.48, 42)<sup>a</sup> | 5.02 (0.40, 65)<sup>d,e</sup> | 7.89 (0.41, 59)<sup>c,f</sup> | 11.08 (0.49, 59)<sup>a,d,f</sup> |

CDR = Clinical Dementia Rating; CVLT = California Verbal Learning Test; FAQ = Functional Activities Questionnaire; MMSE = Mini Mental State Exam; NPI = Neuropsychiatric Inventory; NA = Not Applicable; VOSP = Visual Object and Space Perception Battery. W = White, A = Asian, N = Native American. EMM = estimated marginal means, se = standard error.
Right temporal different from health control at <.05: a
Right temporal different from frontal at <.05: b
Right temporal different from left temporal at <.05: c
Frontal different from health control at <.05: d
Frontal different from left temporal at <.05: e
Left temporal different from health control at <.05: f

**Table 2:** Language and socioemotional profile.

|  | Right Temporal | Frontal | Left Temporal | Healthy Control |
| --- | --- | --- | --- | --- |
| **Language** |  |  |  |  |
| Verbal agility, EMM (se, n), max =5 | 4.94 (0.19, 36) | 4.73 (0.16, 54)<sup>e</sup> | 5.44 (0.16, 57)<sup>e</sup> | 5.25 (0.19, 52) |
| Repetition, EMM (se, n), max =5 | 4.16 (0.19, 36)<sup>c</sup> | 3.84 (0.16, 57)<sup>d</sup> | 3.46 (1.52, 57)<sup>f</sup> | 4.43 (0.19, 59)<sup>c,d,f</sup> |
| WRAT Reading, EMM (se, n), max =70 | 56.64 (3.02, 21)<sup>a</sup> | 54.71 (2.18, 41)<sup>d</sup> | 50.00 (2.26, 38)<sup>f</sup> | 65.22 (2.11, 58)<sup>a,d,f</sup> |
| Apraxia of speech rating, EMM (se, n), max =7 | 0 (0.00, 27) | 0.14 (0.08, 14) | 0.06 (0.04, 49) | 0.0 (0.00, 9) |
| Dysarthria rating, EMM (se, n), max =7 | 0.09 (0.04, 43)<sup>b</sup> | 1.3 (0.09, 40)<sup>b,d,e</sup> | 0.06 (0.04, 50)<sup>e</sup> | 0.14 (0.07, 59)<sup>d</sup> |
| Syntax Comprehension, EMM (se, n), max =5 | 4.43 (0.14, 36) | 3.97 (0.12, 56)<sup>e</sup> | 4.48 (0.12, 56)<sup>e</sup> | 4.38 (0.14, 52) |
| Lexical fluency – # in 60", EMM (se, n) | 8.53 (0.64, 43)<sup>ab</sup> | 6.16 (0.61, 69)<sup>b,de</sup> | 8.51 (0.53, 66)<sup>c,f</sup> | 14.09 (0.81, 35)<sup>a,d,f</sup> |
| **Verbal Semantics** |  |  |  |  |
| BNT, EMM (se, n), max = 15 | 6.89 (0.43, 33)<sup>a,b,c</sup> | 12.02 (0.35, 70)<sup>b,e</sup> | 5.18 (0.36, 66)<sup>c,e,f</sup> | 12.99 (0.46, 57)<sup>a,f</sup> |
| Verbal fluency – semantic – # in 60 seconds, EMM (se, n) | 9.21 (0.76, 43)<sup>a</sup> | 10.78 (0.74, 69)<sup>d</sup> | 8.99 (0.64, 66)<sup>f</sup> | 21.62 (0.78, 35)<sup>a,d,f</sup> |
| PPVT, EMM (se, n), max =16 | 9.56 (0.43, 40)<sup>a,b</sup> | 13.62 (0.45, 65)<sup>b,e</sup> | 9.53 (0.39, 53)<sup>c,f</sup> | 13.97 (0.46, 53)<sup>a,f</sup> |
| Pyramids and Palm Trees Percent, Pictures, EMM (se, n), max =1 | 0.81 (0.02, 27)<sup>a,b</sup> | 0.90 (0.01, 49)<sup>b</sup> | 0.86 (.01, 53) | 0.90 (.01, 30)<sup>a</sup> |
| **Face perception** |  |  |  |  |
| CATS Face Matching, EMM (se, n), max =12 | 11.24 (0.20, 31) | 10.66 (0.18, 42)<sup>d,e</sup> | 11.79 (0.17, 41)<sup>e</sup> | 11.55 (0.18, 52)<sup>d</sup> |
| **Person-specific knowledge** |  |  |  |  |
| Famous Faces Naming, EMM (se, n), max =20 | 1.26 (0.86, 15)<sup>a,b</sup> | 6.88 (1.19, 9)<sup>b,de</sup> | 1.75 (1.00, 17)<sup>e</sup> | 12.82 (0.87, 26)<sup>a,d,f</sup> |
| Famous Faces Familiarity, EMM (se, n), max =20 | 6.85 (0.88, 24)<sup>a,b,c</sup> | 17.48 (1.70, 7)<sup>b</sup> | 14.80 (0.92, 32)<sup>c</sup> | 14.19 (1.10, 26)<sup>a</sup> |
| Famous Faces Semantic Association, EMM (se, n), max =20 | 5.37 (1.13, 12)<sup>a,b,c</sup> | 16.19 (1.2, 8)<sup>b,e</sup> | 11.75 (1.37, 10)<sup>c,e</sup> | 14.71 (0.96, 26)<sup>a</sup> |
| Famous Faces Name Familiarity, EMM (se, n), max =16 | 2.80 (0.78, 14) | 9.80 (1.06, 10) | 2.92 (0.78, 10) | 11.91 (0.94, 29) |
| **Social Function and Emotion** |  |  |  |  |
| CATS Affect Matching, EMM (se, n), max =16 | 9.11 (0.42, 35)<sup>a,c</sup> | 9.67 (0.39, 43)<sup>d,e</sup> | 12.82 (0.41, 40)<sup>c,e</sup> | 12.82 (0.46, 52)<sup>a,d</sup> |
| TASIT Emotion Evaluation Test, EMM (se, n), max =14 | 6.46 (0.48, 27)<sup>a</sup> | 7.77 (0.40, 45)<sup>d</sup> | 8.19 (0.42, 40)<sup>f</sup> | 10.89 (0.40, 57)<sup>a,d,f</sup> |
| TASIT SI-M Sincere, EMM (se, n), max =20 | 15.99 (0.69, 24)<sup>b</sup> | 13.42 (0.52, 51)<sup>b,de</sup> | 17.16 (0.55, 42)<sup>e</sup> | 16.64 (0.55, 58)<sup>d</sup> |
| TASIT SI-M Sarcastic, EMM (se, n), max =20 | 4.74 (0.85, 24)<sup>a,b,c</sup> | 13.49 (0.65, 51)<sup>b,de</sup> | 9.80 (0.67, 42)<sup>c,e,f</sup> | 17.60 (0.68, 58)<sup>a,d,f</sup> |
| IRI-Empathetic Concern, EMM (se, n), max =24 | 16.09 (1.41, 44)<sup>a,c</sup> | 14.94 (1.12, 79)<sup>d,e</sup> | 21.87 (1.14, 75)<sup>c,e,f</sup> | 27.41 (1.56, 54)<sup>a,d,f</sup> |
| IRI-Perspective Taking, EMM (se, n), max =24 | 10.77 (1.10, 44)<sup>a,c</sup> | 10.22 (0.87, 79)<sup>d,e</sup> | 14.63 (0.89, 75)<sup>c,e,f</sup> | 22.86 (1.22, 54)<sup>a,d,f</sup> |
| Emotional Theory of Mind, EMM (se, n), max =16 | 12.25 (0.46, 9)<sup>a</sup> | 12.44 (0.29, 23)<sup>d,e</sup> | 13.86 (0.36, 17)<sup>e</sup> | 14.62 (0.35, 20)<sup>a,d</sup> |
| Cognitive Theory of Mind, EMM (se, n), max =16 | 14.79 (0.58, 15)<sup>b</sup> | 12.04 (0.44, 30)<sup>b,de</sup> | 15.07 (0.44, 33)<sup>d</sup> | 15.07 (0.35, 59)<sup>e</sup> |
| Behavioral Inhibition BIS-Total, EMM (se, n), max =24 | 17.39 (0.73, 24) | 17.49 (0.55, 54)<sup>e</sup> | 19.85 (0.59, 41)<sup>e,f</sup> | 15.95 (0.61, 54)<sup>f</sup> |
| Behavioral Activation BAS-Drive, EMM (se, n), max =24 | 8.56 (0.56, 25) | 10.21 (0.42, 50) | 9.99 (0.44, 38) | 10.40 (0.47, 54) |
| Behavioral Activation BAS-Fun, EMM (se, n), max =24 | 7.54 (0.49, 25)<sup>a,b</sup> | 9.47 (0.39, 50)<sup>b</sup> | 8.69 (0.42, 36) | 9.50 (0.42, 53)<sup>a</sup> |
| Behavioral Activation BAS-Reward, EMM (se, n), max =24 | 13.13 (0.52, 25)<sup>a,c</sup> | 13.59 (0.42, 51)<sup>e</sup> | 15.69 (0.44, 38)<sup>c,e</sup> | 15.08 (0.45, 54)<sup>a</sup> |
| IAS - Current warmth, EMM (se, n) | 37.59 (3.02, 13)<sup>a,c</sup> | 38.86 (1.95, 36)<sup>d,e</sup> | 46.56 (1.84, 44)<sup>c,e</sup> | 47.89 (2.12, 44)<sup>a,d</sup> |
| IAS - Current dominance, EMM (se, n) | 37.24 (2.59, 13) | 30.86 (1.68, 36)<sup>d,e</sup> | 37.76 (1.58, 44)<sup>e</sup> | 42.21 (1.82, 44)<sup>d</sup> |
| IAS - Current Coldness, EMM (se, n) | 29.53 (2.27, 13)<sup>a,c</sup> | 24.77 (1.71, 36)<sup>d</sup> | 18.88 (1.71, 44)<sup>c</sup> | 13.72 (1.83, 44)<sup>a,d</sup> |
| Self-Monitoring RSMS, EMM (se, n), max =65 | 36.57 (2.32, 28)<sup>a,c</sup> | 38.68 (1.78, 55)<sup>d,e</sup> | 45.14 (1.86, 49)<sup>c,e,f</sup> | 59.05 (2.24, 47)<sup>a,d,f</sup> |
| Depression GDS, EMM (se, n), max =30 | 7.56 (0.94, 37)<sup>a</sup> | 8.88 (.87, 54)<sup>d</sup> | 8.54 (.83, 50)<sup>f</sup> | 3.38 (0.92, 58)<sup>a,d,f</sup> |

BNT = Boston Naming Test; PPVT = Peabody Picture Vocabulary Test; CATS = Comprehensive Affect Testing System; TASIT = The Awareness of Social Inference Test; EET = Emotion Evaluation Test; IRI = Interpersonal Reactivity Index; EC = Empathic Concern; PT = Perspective Taking; IAS = Interpersonal Adjective Scales; RSMS = Revised Self-Monitoring Scale. GDS = Geriatric Depression Scale. \* = Autopsy Group and Living Group are statistically different $p < .05$ . W = White, A = Asian, N = Native American. EMM = estimated marginal means, se = standard error.
Right temporal different from health control at $<.05$ : a
Right temporal different from frontal at $<.05$ : b
Right temporal different from left temporal at $<.05$ : c
Frontal different from health control at $<.05$ : d
Frontal different from left temporal at $<.05$ : e
Left temporal different from health control at $<.05$ : f

We assessed multiple domains of socioemotional functioning with a battery of task-based measures. Visual face perception was evaluated with the identity-matching subtest of the Comprehensive Affect Testing System (CATS), in which patients determined whether pairs of neutral faces were from the same person or different people.^61^ The ability to label emotional facial expressions with words was tested with the CATS emotion identification task, in which patients chose the emotion term that matched the facial expression depicted in a photograph from a list of multiple choice options.^62^ On the abbreviated version of the Emotion Evaluation Test (EET) from The Awareness of Social Inference Test (TASIT), patients identified the target emotion from a list of multiple choice options that were displayed by actors in short video clips. On the TASIT Social Inference–Minimal Test (SIM), patients were asked to detect sarcasm of actors in videos through interpretation of social cues including prosody, facial expression, and gesture. Theory of mind (ToM)—the ability to infer the thoughts, emotions, and intentions of others—was tested in cognitive (i.e., the ability to identify first and second order object knowledge of actors in videos) and emotional modalities (i.e., the ability to identify first and second order emotion knowledge of actors in videos) using the UCSF theory of mind Test.^63^ Person-specific semantic knowledge was evaluated using the UCSF Famous Faces Naming test (a free response task in which patients named photographs of famous people’s faces), Semantic Famous Face Association test (patients matched famous faces based on their professions), Semantic Famous Name Association test (patients matched written names of famous people based on their professions), and Semantic Famous Face Recognition test (patients chose the famous face among four faces)^11,64^ Further socioemotional testing details are found in the Supplementary material.

Informant-based measures were also obtained to assess patients’ socioemotional behavior in everyday life. Informants rated patients’ current cognitive empathy (i.e., perspective taking) and emotional empathy (i.e., empathic concern) using the Interpersonal Reactivity Index (IRI).^65^ Sensitivity and responsiveness to others’ subtle emotional expressions were rated by informants using the Revised Self-Monitoring Scale (RSMS).^66^ Interpersonal coldness, warmth, and dominance, areas of personality known to be affected in FTD, were evaluated with informant ratings on the Interpersonal Adjective Scales (IAS).^67^ Behavioral inhibition (i.e., behaviors associated with response avoidance and sensitivity to threat) and behavioral activation (i.e., behaviors associated with approach motivation including reward responsiveness, drive, and fun-seeking) systems were evaluated with informant ratings on the Behavioral Inhibition System/Behavioral Activation System (BIS/BAS) questionnaire.^68^

### Structural neuroimaging analyses

We processed structural T1-weighted images, as previously described.^69,70^ W-score maps (W-maps) were generated by comparing each patient’s gray matter maps to 534 neurologically healthy older controls from the MAC Hillblom Healthy Aging Network (age range 44-99 years, M±SD: 68·7±9·1; 220 male/302 female), adjusted for age, sex, total intracranial volume, and magnet field strength. Mean W-score values were extracted for each region of interest (ROI) in the probabilistic Desikan atlas. W-scores have a mean value of 0 and a standard deviation (SD) of 1; values of +1.65 and -1.65 correspond to the 95th and 5th percentiles and indicate regions with larger and smaller gray matter volume compared to the normative sample, respectively.

Patients were included in the rATL-predominant degeneration group if their lowest three W-scores were in right temporal regions, and they had relative preservations of the frontal lobes based on an atrophy index described as follows. For each patient with rATL maximum atrophy, we calculated the mean W-score of all frontal lobe ROIs and the mean W-score of all right temporal ROIs and computed a proportion with the following index: *right temporal index = mean whole frontal w-score/mean right temporal w-score*. The rATL-predominant degeneration patients who had an index < 0.50 were included in this study (n = 46) (Fig. 2 and Supplementary Table 1). A similar approach was used to select the comparison groups. Patients were included in the frontal-predominant group if their lowest three W-scores were in the frontal regions, and they had relative preservation of the right temporal regions based on an atrophy index (mean frontal W-map score / mean right temporal W-map score > 0.50). Patients were included in the lATL-predominant group if their lowest three W-map scores were in the left temporal regions, and they had relative preservation of the frontal lobes based on atrophy index (mean frontal W-map score / mean left temporal W-map score < 0.50). We implemented this index for the lATL instead of a right / left temporal laterality index in order to match the rATL and lATL patients based on their degree of accompanying frontal involvement.

**Figure 1:**
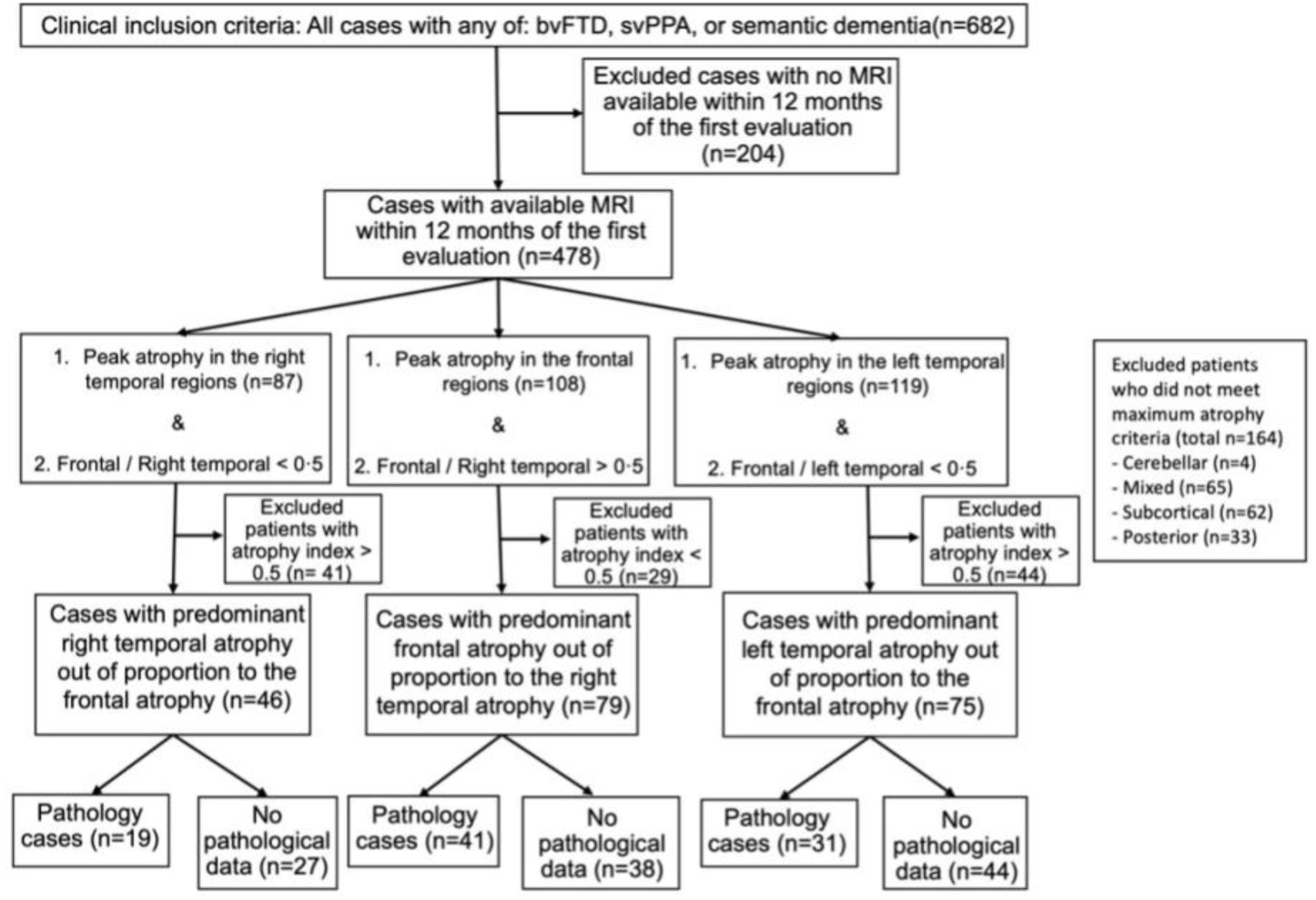
Patient selection. We searched the University of California, San Francisco Memory and Aging Center database. The first inclusion criteria was the clinical diagnosis, we included all participants who received a clinical diagnosis of behavioral variant frontotemporal dementia or semantic variant primary progressive aphasia. We then excluded all patients who did not have a brain MRI within one year of the first research evaluation. Next, we included participants who had peak atrophy in either the right temporal lobe, frontal lobe, or left temporal lobe on a brain MRI W-score map and showed predominant atrophy in their respective lobe based on an atrophy index. bvFTD = behavioral variant frontotemporal dementia spectrum disorders; svPPA = semantic variant primary progressive aphasia, posterior = parietal or occipital lobes.

**Figure 2:**
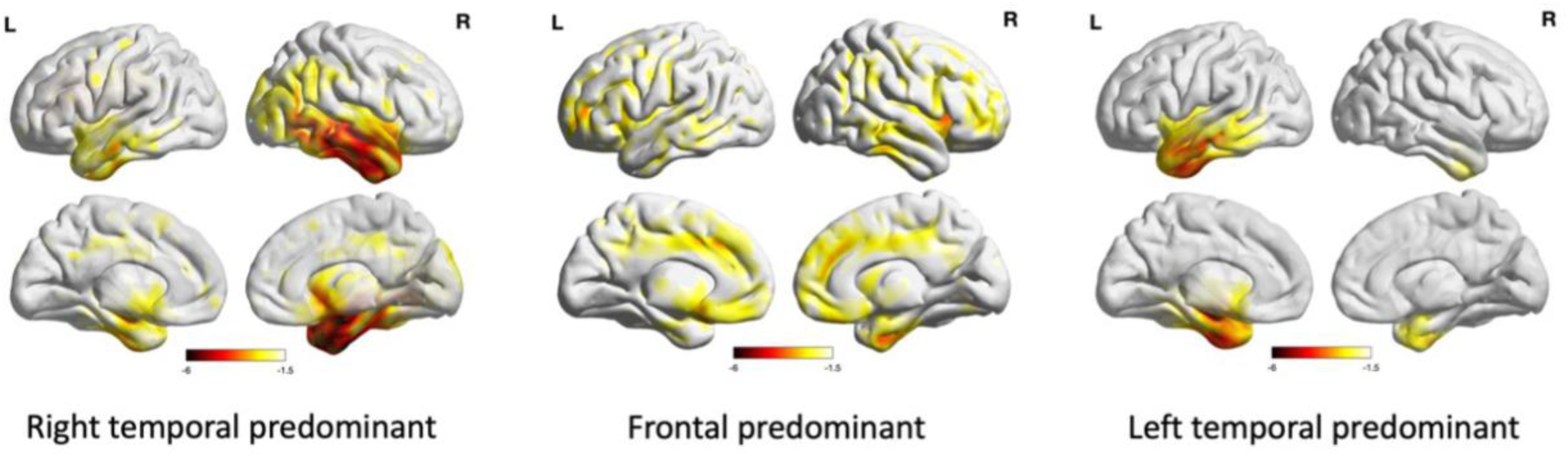
Neuroimaging in right temporal-, left temporal-, and frontal-predominant neurodegeneration. Lateral and mesial views. Predominant right temporal, left temporal, or frontal atrophy was used as part of the inclusion criteria based on a data-driven neuroimaging approach. The right temporal predominant group exhibited maximum atrophy in the right temporal lobe more than the left anterior temporal lobe with involvement of the right more than left insula, right caudate, and right more than left subgenual anterior cingulate cortex. Notably there is sparing of the frontal, parietal, and occipital lobes. The left temporal predominant group shows maximum atrophy in the left temporal lobe more than the right anterior temporal lobe with involvement of the left more than right insula, left caudate, and left subgenual anterior cingulate cortex. Further, there is sparing of the frontal, parietal, and occipital lobes. The frontal group shows bilateral lateral and mesial frontal and left temporal volume loss but relative sparing of the right temporal lobe.

Patients were excluded if they did not meet either the predominant atrophy or the atrophy index requirements. Patients were excluded if their lowest W-scores were not in the rATL, frontal lobe, or lATL (n = 164; four cerebellar, 65 mixed (i.e., lowest three W-scores were in different lobes), 62 subcortical, and 33 posterior (i.e., parietal or occipital lobes)), or if their greatest atrophy was in the rATL, lATL, or frontal lobes but did not meet the atrophy index inclusion threshold (total n = 114; 41 with maximum atrophy in rATL but frontal / right temporal > 0.5, 29 with maximum atrophy in the frontal lobe but frontal / right temporal < 0.5, and 44 with maximum atrophy in the lATL but frontal / left temporal > 0.5).

Recognizing that each of the groups included in this study did not show atrophy merely in one isolated brain region (for instance, patients typically have bilateral ATL volume loss by the time they present for imaging evaluation), we qualify our descriptions by using the term “predominant” to refer to the patient group with maximum atrophy in the one region that is out of proportion to the other regions. Thus, we acknowledge that the brain atrophy pattern of our “rATL-predominant” group also includes frontal and left temporal regions to varying degrees, but these patients unequivocally present with maximum atrophy in the rATL. Similarly, the “lATL-predominant” refers to the patient group with maximum atrophy in the lATL that is out of proportion to the frontal and rATL atrophy. The term “frontal-predominant’’, likewise, is used to refer to the patient group with maximum atrophy in the frontal lobes that is out of proportion to the ATL atrophy. The atrophy of each group as shown in Figure 2 extends beyond the regions of maximum atrophy and as such symptoms could be due to atrophy in the other regions involved or multiple parts of the connected networks”

### Genetic and neuropathological data

Participants were screened for the following genetic mutations: *PGRN, MAPT, TARDBP, C9orf72, APP, PSEN1, PSEN2, FUS*, and *APOE*. In the patients who underwent autopsy, brains were processed and analyzed according to the UCSF Neurodegenerative Disease Brain Bank protocol.^71^ In short, eight micro-thick formalin-fixed paraffin-embedded tissue sections from 23 tissue blocks were cut to represent 27 regions of interest. All blocks underwent routine hematoxylin and eosin staining, and subsets underwent immunohistochemistry for hyperphosphorylated tau, amyloid-β, TDP-43, alpha-synuclein, and 3R-tau antibodies. Neuropathological diagnoses were based on consensus criteria.^72–74^

### Statistical analysis

Tests of normality for all continuous data were conducted with the Shapiro-Wilk test. Homogeneity of variance was tested by Levene’s test. Statistical differences in the frequency of categorical variables across groups such as clinical symptoms and APOE genotype were performed with the Chi-Square test. Means of demographic measures (Table 1) were compared across groups with the ANOVA test. Means of functional, neuropsychological, language, and socioemotional measures (Tables 1 and Table 2) were compared with the ANCOVA test correcting for age, sex, and disease severity as measured by the Mini-Mental State Exam (MMSE). Because of the unequal sample sizes and unequal group variances, pairwise post hoc comparisons were done with estimated marginal means and Bonferroni-Sidak adjusted probabilities to correct for multiple comparisons, with *p* < 0.05 set as the threshold for statistical significance. Data analysis was performed with SPSS (version 27, SPSS/IBM, Chicago, IL). Table 1 and Table 2 show estimated marginal means, standard errors, and statistical significance after correcting for age, sex, and disease severity as measured by MMSE. When using ANCOVA and estimated marginal means for post hoc between groups analysis, the individual datapoints cannot be graphically plotted, for visualization purposes we show in Figure 4 the uncorrected datapoints, means, and standard deviations for key socioemotional measures.

### Data availability

The data for this study is available upon request. The sensitive nature of patients’ data and the institutional ethics protocols in place at the time these patients gave informed consent do not permit open data sharing. The clinical and neuroimaging data used in the current paper are available from the senior author (MLGT), upon formal request indicating name and affiliation of the researcher as well as a brief description of the use that will be done of the data. All requests will undergo UCSF-regulated procedure thus requiring submission of a Material Transfer Agreement (MTA). No commercial use would be approved.

## Results

### Demographic features

Table 1 shows the demographic information. Although the sex distribution was not different between healthy control and the patient groups, the healthy controls were older than all of the patient groups. Patients in all cohorts were highly educated with an average over 15.5 years of education. In the rATL-predominant group, 91% were White and 9% were Asians, a proportion that was not different from other groups. In the rATL cohort (n = 46), the average age of onset (60.2 years and sd = 6.8 years). In the rATL-predominant group, 52% of the patients were men and 15% were non-right-handed. On average, rATL-predominant patients were in the mild to moderate range of disease severity; at the first research visit, the average MMSE score (25.7/30; sd = 5.2) was higher than the other disease groups. The CDR for the rATL group (average score 0.9/3; sd = 0.5) was lower than the lATL-predominant group but not different from the frontal-predominant group. We used age, sex, and MMSE as confounds in all later analyses.

### Diagnostic criteria and clinical symptom chronology

During the first three years of the illness, only a minority of patients in the rATL-predominant group met diagnostic criteria for Neary-FTD (13%), Neary-semantic dementia (9%), bvFTD (27%), or svPPA (13%). Approximately one third of the group had verbal svPPA features (i.e., impaired confrontational naming and object knowledge) but did not meet the general criteria for PPA (36%) because aphasia was not the initial and predominant symptom. At the time of the first research evaluation at the MAC (average 5.3 years after disease onset), these percentages were higher: Neary-FTD (52%), Neary-semantic dementia (11%), bvFTD (83%), svPPA (16%), and semantic variant features (78%) (Supplemental Table 2).

The clinical histories revealed that, when combining all symptoms that emerged during the first three years of the illness, the most common symptoms for patients with rATL-predominant degeneration were loss of empathy (27%), loss of person-specific semantic knowledge (23%), complex compulsions and rigid thought process (18%), and loss of verbal semantic knowledge (13%) (Fig. 3). The sequence of the first two symptoms in rATL-predominant patients is shown in Supplemental table 3.

**Figure 3:**
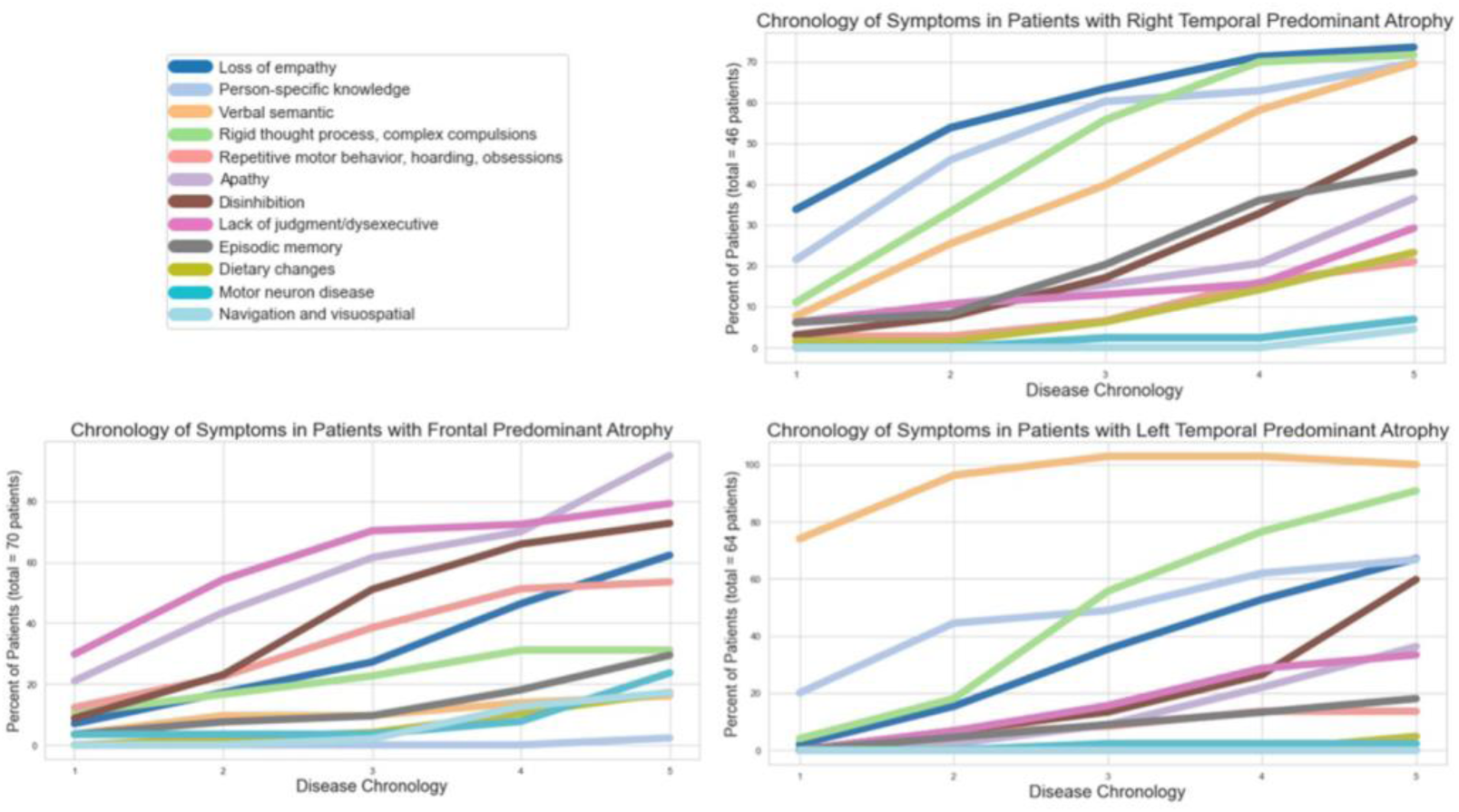
Chronology of symptoms. Left upper corner shows the symptom legend. Right upper corner shows symptoms chronology in the right temporal-predominant group; the most common early symptoms in this group are loss of empathy, loss of person-specific knowledge, and rigid thought process and complex compulsion. The left lower corner shows symptom chronology in the frontal-predominant group; the most common early symptoms in this group are lack of judgment/dysexecutive symptoms, apathy, and disinhibition. The right lower corner shows symptom chronology in the left temporal-predominant group; the most common early symptoms in this group are verbal semantic loss, loss of person-specific knowledge, and rigid thought process and complex compulsion.

**Figure 4:**
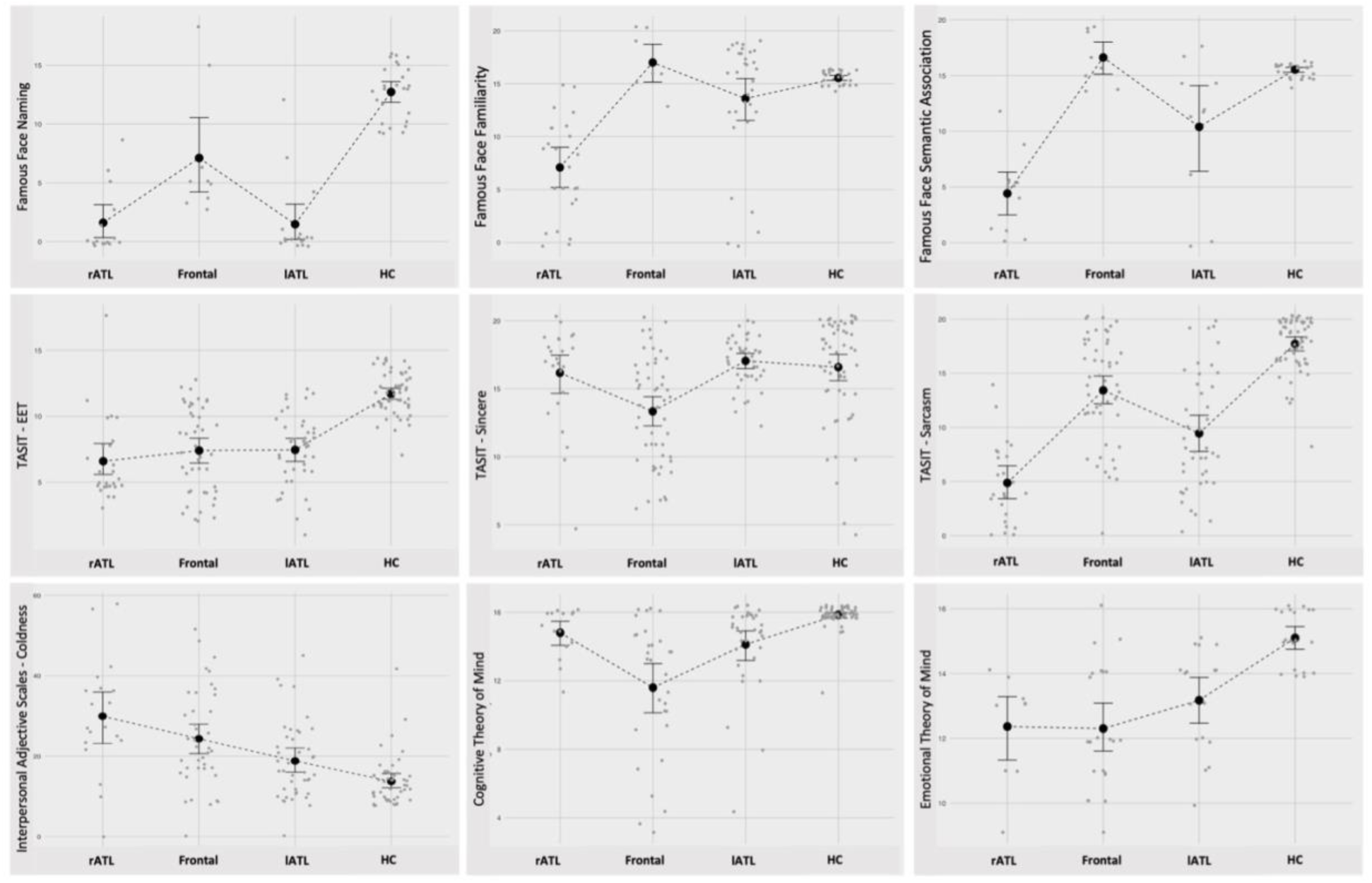
Socioemotional and Neuropsychological characteristics. The figure shows the results of the main socioemotional tests that can help distinguish right temporal-from frontal-predominant patients. More details can be found in Table 2. Although all disease groups had difficulties with famous faces naming, only right temporal, and a lesser degree left temporal-predominant patients, had difficulties on Famous Face Familiarity and Semantic Association. Although all disease groups showed impaired simple and complex social cues recognition on the TASIT-EET and TASIT-Sarcasm, only the frontal predominant group showed impairment on the control cognitive task, TASIT-Sincere. Right temporal-predominant patients showed significantly worse performance on the complex social cue, TASIT-Sarcasm, compared to the frontal predominant group. The right temporal-predominant group showed increased coldness compared to the frontal predominant group. The right and left temporal-predominant groups had difficulty with the emotional theory of mind but not with the cognitive theory of mind task. In contrast to the frontal-predominant group which demonstrated impairment in both cognitive and emotional theory of mind.

Caregiver-reported examples of loss of empathy included decline in the ability to understand and respond to others’ emotions and needs (e.g., not consoling a family member who lost a parent or was diagnosed with a terminal illness, making tactless comments in a funeral, asking a crying child why their eyes were watering, and becoming more self-centered). In our experience, often, loss of empathy toward others can be interpreted by caregivers as selfishness. Examples of loss of person-specific semantic knowledge included not recognizing familiar people by the face or voice, not recalling biographical information of a famous person, and not knowing patients’ own relationship to familiar people. Examples of complex compulsions and rigid thought process included adherence to rigid time schedules; dogmatism; hypergraphia; hypochondriasis; restricted color, clothing, diet, game, or puzzle preferences. Less commonly, patients exhibited simple repetitive motor or speech behaviors or hoarding behaviors. Examples of loss of verbal semantic knowledge included difficulty understanding word meaning or recognizing objects.

When rATL-predominant patients had both person-specific and verbal semantic knowledge loss (32 patients [69%]), the person-specific semantic knowledge symptoms were reported to precede the verbal semantic complaints in 24 patients (75%). Five patients (10%) had person-specific semantic knowledge symptoms without verbal semantic complaints, and six patients (14%) had verbal semantic complaints without person-specific semantic knowledge complaints. Only three patients (6%) had neither person-specific nor verbal semantic knowledge complaints.

While these initial symptoms in the rATL-predominant group emerged within the first three years of disease onset, additional symptoms (fifth, sixth, and beyond) arose as the disease progressed. Four years after disease onset, common symptoms included apathy and disinhibition. For these two symptoms, differences in reporting created ambiguity. Apathy was explicitly documented in the medical history as a clinical complaint of 11 patients, whereas on the NPI (Supplementary Table 5), apathy was noted in 39 patients, indicating a discrepancy between what caregivers report during the interview with the behavioral neurologist and when answering the NPI questions. Interestingly, the item of apathy on the NPI appeared mainly in the context of affective apathy questions rather than cognitive inertia or autoactivation/behavioral apathy and thus these behaviors could also be interpreted as loss of empathy. This potentially explains the discrepancy between clinical history and NPI with regards to apathy reporting and highlights the need for incorporating loss of empathy questions into the NPI. On history, disinhibition was reported in 23 patients, whereas on the NPI it was coded in 36 patients. In the majority of patients, disinhibition appeared as insensitivity to social context rather than as an impulse-control deficit, for instance making funny comments in a funeral rather than approaching strangers or engaging in dangerous activities. By history, episodic memory impairment, executive symptoms, dietary changes, motor neuron disease, and problems navigating were less frequent and happened later in the disease course. With regards to the less commonly reported symptoms in the rATL-predominant group, five patients (11%) had loss of sexual desire, two as an early symptom. Irritability was reported in eight patients (17%) and as an early symptom only in three patients (6%). Increased eating (7 patients, 8%) did not reach the degree of binge eating, oral exploration or consumption of inedible objects. Sleep changes, increased or decreased sleep, happened in five cases (10%), three of which were in the first year of disease onset.

In comparison, the early symptoms in the frontal-predominant group were lack of judgment/dysexecutive symptoms (24%), apathy (21%), and disinhibition (17%), as shown in Fig 3 and Supplementary Table 4. In the lATL-predominant patients, the early symptoms were verbal semantic knowledge loss (36%), person-specific knowledge deficits (16%), and rigid thought process (18%). Loss of empathy occurred significantly more often in rATL-compared to frontal- and lATL-predominant patients (*χ*^*2*^ = 22, *p <*.001 and 11.2 *p <*.001, respectively). Deficits in person-specific knowledge were significantly more common in rATL-than frontal-predominant patients (*χ*^*2*^ = 56.1, p < .001) but not lATL-predominant patients (*χ*^*2*^ = 3.32, *p* < .68). Similarly, complex compulsions and rigid thought process was significantly more frequent in rATL-compared to frontal-, but not lATL-predominant groups (*χ*^*2*^ = 19.54, *p* < .001 and *χ*^*2*^ = 1.03, *p* = 0.3, respectively). In contrast apathy, disinhibition, and lack of judgment/dysexecutive symptoms were significantly more common in frontal- than rATL- and lATL-predominant patients (*χ*^*2*^ = 11.5, *p* < .001, *χ*^*2*^ = 5.2, p < .02, *χ*^*2*^ = 18.8, *p* < .001, respectively).

### Functional, cognitive, and behavioral results

Table 1, Table 2, and Figure 4 show the neuropsychological and socioemotional results. Neuropsychological testing demonstrated that, at presentation to the MAC, patients with rATL-predominant degeneration had severe impairment in both verbal semantic knowledge (on the BNT and PPVT) and nonverbal (visual) semantic knowledge (on the PPT-P). They also had deficits in verbal fluency, with more significant impairment in semantic than in lexical fluency, and on tests of executive functioning. Episodic memory was impaired, and visuospatial processing was intact.

On tests of socioemotional functioning, rATL-predominant degeneration patients had severe deficits in multiple domains. On the CATS, a static face perception test, although they had no difficulty with face identity-matching, their emotion labeling was impaired, suggesting a deficit in emotion recognition. Patients also had difficulty labeling the emotions of others in videos (TASIT-EET) and understanding paralinguistic cues (TASIT-SIM-M). On tests of Theory of Mind, patients had normal cognitive theory of mind scores but impaired emotional theory of mind scores, indicating poor comprehension of others’ emotional, but not cognitive, states. On the Famous Faces test, rATL-predominant degeneration patients could not identify the faces, names, or occupations, of famous people, indicating loss of person-specific semantic knowledge rather than prosopagnosia. On informant-based measures, rATL degeneration patients had abnormal scores on multiple measures of behavior and personality. Patients had very low emotional empathy (IRI Empathic Concern), cognitive empathy (IRI Perspective Taking) and socioemotional sensitivity (RSMS). On a personality inventory (IAS), informants rated patients as having low levels of interpersonal warmth and increased interpersonal coldness yet preserved interpersonal dominance.

Although emotion processing was disrupted in frontal-, rATL-, and, to a lesser degree, lATL-predominant patients (as measured by IRI-ET, IRI-PT, and RSMS), the groups differed in their specific constellations of social and behavioral deficits. While the frontal-predominant patients were impaired on both cognitive and emotional measures, the rATL- and lATL-predominant patients generally showed prominent deficits on the emotional, but not the cognitive, components of socioemotional tasks. Specifically, rATL- and lATL-predominant patients showed preserved cognitive theory of mind but impaired emotional theory of mind, whereas frontal-predominant patients showed impairment on both cognitive and emotional theory of mind (Figure 4 and Table 2). Similarly, rATL-predominant patients scored within normal limits, on the TASIT-Sincere task (a cognitive control task that assesses simple comprehension) but below expectations on the TASIT – EET (an emotion naming task) and the TASIT – Simple Sarcasm subscale (a test of paralinguistic cue detection). In contrast, the frontal-predominant group scored below expectations on all three TASIT subsets, suggesting both emotional and cognitive deficits. On informant-based personality measures, the rATL-predominant patients showed increased coldness but preserved dominance, whereas the frontal-predominant patients showed increased coldness (to a lesser degree than rATL-predominant patients) but reduced dominance. Furthermore, rATL-predominant patients showed reduction in both their activation and inhibition systems on the BIS/BAS. Reduced reward sensitivity was associated with reduced drive and fun-seeking in rATL-predominant patients, whereas in the frontal-predominant patients reduced reward sensitivity was associated with higher drive and fun-seeking (Fig 4, Table 2). This incongruence in the frontal-predominant group is consistent with the higher rates of impulsivity, such as making sexual comments, in this group as shown on the NPI (Supplemental table 5).

With regard to face processing and person-specific knowledge, while all disease groups had difficulty with Famous Faces Naming, only rATL-predominant patients (and to a lesser degree lATL-predominant patients) had impaired scores on the Famous Faces Familiarity and Semantic Association tests (Figure 4 and Table 2). On the CATS Face and Affect Matching, whereas the frontal-predominant patients were impaired on both tests, the rATL-predominant patients had preserved face matching but impaired affect matching. The lATL-predominant patients, in contrast, had intact scores on the Face and Affect Matching tests (Figure 4 and Table 2). The deficits in the rATL group did not appear to be due to broader deficits in visuospatial functioning as only the frontal-predominant patients demonstrated deficits in this area (Benson Figure Copy and Visual Object and Space Perception).

With regard to verbal semantics, both rATL- and lATL-predominant patients had greater deficits on the BNT than the frontal-predominant group. On episodic memory testing, rATL- and lATL-predominant patients showed worse verbal and visual memory impairment compared to the frontal-predominant group (Fig 4 and Table 2). On executive function tests, rATL- and lATL-predominant patients showed better executive function performance compared to the frontal-predominant group (Fig 4 and Table 1).

### Genetic and pathology results

Pathology and genetic results are presented in Tables 3 and 4. Only two of the rATL-predominant patients had a genetic mutation, (one had a *MAPT* mutation, and one had a possibly pathogenic *TARDBP* mutation). Seventeen of the frontal-predominant patients (14 *C9orf72* and three *GRN*) and five lATL-predominant patients (three *MAPT* and two *C9orf72*) had genetic mutations. *APOE* data were available in 40 of the rATL-predominant patients (55% E3/E3; 22% E3/E4; 18% E2/E3). No differences in *APOE* genotypes were found between subgroups with available *APOE data* (Supplementary table 7).

See Table 3 and Supplementary Table 6 for the pathology results. Most of the rATL-predominant patients with available autopsy data had FTLD-TDP type C pathology (68%). When considering all types of FTLD-TDP cases, regardless of the neuropathological subtype, the percentage increased (84%). Three patients had FTLD-tau (two FTLD-tau Pick’s type, and one patient had FTLD-tau unclassifiable 4R tauopathy). In the rATL-predominant group with autopsy data, three patients did not have loss of semantic knowledge on either history or testing, and, interestingly, none of these three cases had FTLD-TDP type C (two had FTLD-tau Pick’s type, and one had FTLD-TDP type B). In the lATL-predominant group, there was also large proportion of patients with TDP-43 pathology, in general, and TDP-43 type C, specifically. This is in contrast to the frontal-predominant group, which showed more heterogeneity in its underlying pathology (51% tauopathy, 22% FTLD-TDP type B, 12% FTLD-TDP type A, and 2% FTLD-TDP type C) (Table 3).

**Table 3:**
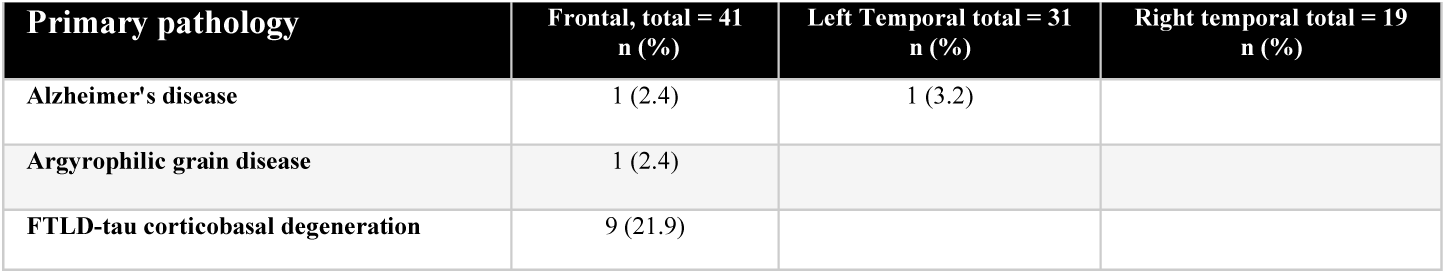
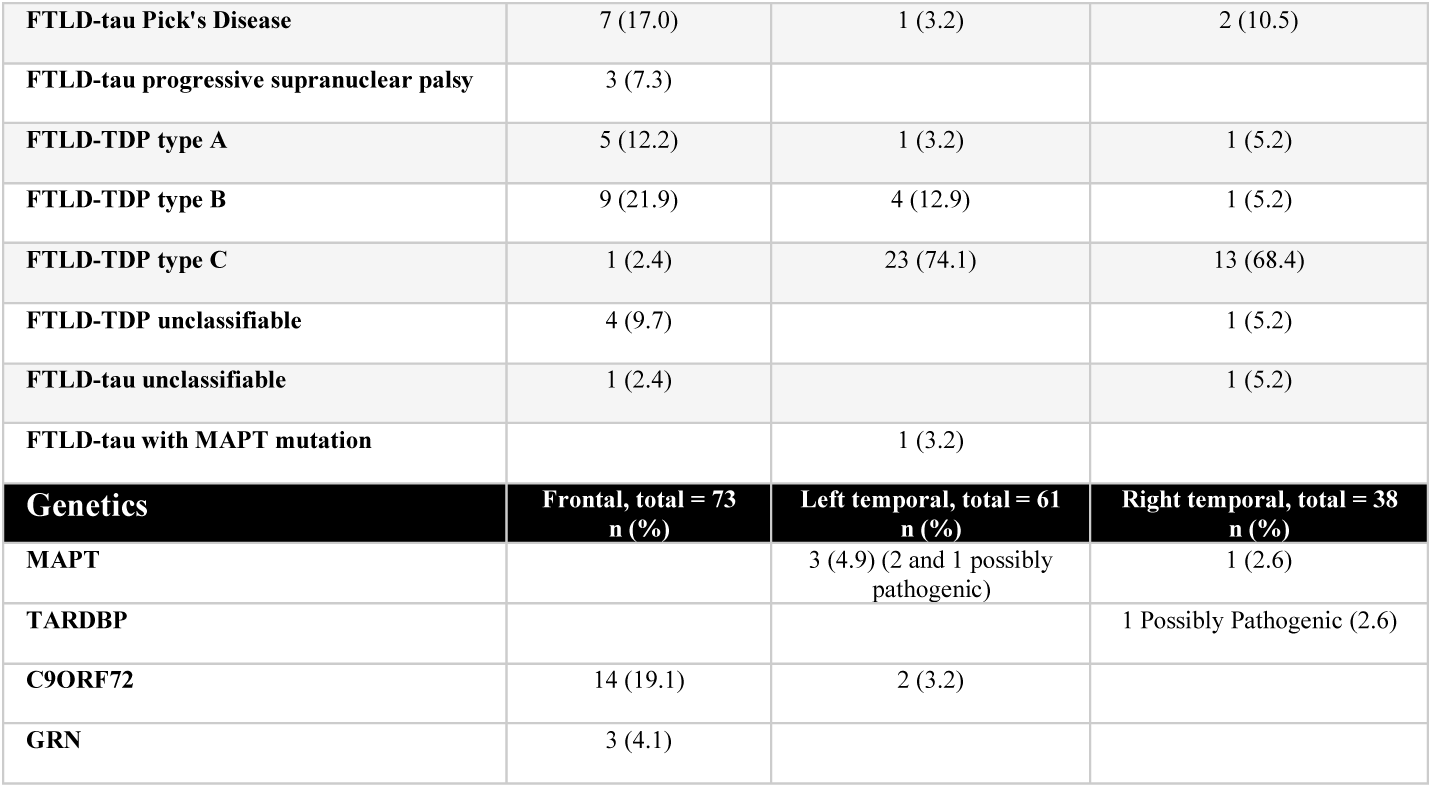
Primary pathology and genetics:

| Primary pathology | Frontal, total = 41<br>n (%) | Left Temporal total = 31<br>n (%) | Right temporal total = 19<br>n (%) |
| --- | --- | --- | --- |
| Alzheimer's disease | 1 (2.4) | 1 (3.2) |  |
| Argyrophilic grain disease | 1 (2.4) |  |  |
| FTLD-tau corticobasal degeneration | 9 (21.9) |  |  |
| FTLD-tau Pick's Disease | 7 (17.0) | 1 (3.2) | 2 (10.5) |
| FTLD-tau progressive supranuclear palsy | 3 (7.3) |  |  |
| FTLD-TDP type A | 5 (12.2) | 1 (3.2) | 1 (5.2) |
| FTLD-TDP type B | 9 (21.9) | 4 (12.9) | 1 (5.2) |
| FTLD-TDP type C | 1 (2.4) | 23 (74.1) | 13 (68.4) |
| FTLD-TDP unclassifiable | 4 (9.7) |  | 1 (5.2) |
| FTLD-tau unclassifiable | 1 (2.4) |  | 1 (5.2) |
| FTLD-tau with MAPT mutation |  | 1 (3.2) |  |
| **Genetics** | **Frontal, total = 73**<br>**n (%)** | **Left temporal, total = 61**<br>**n (%)** | **Right temporal, total = 38**<br>**n (%)** |
| MAPT |  | 3 (4.9) (2 and 1 possibly<br>pathogenic) | 1 (2.6) |
| TARDBP |  |  | 1 Possibly Pathogenic (2.6) |
| C9ORF72 | 14 (19.1) | 2 (3.2) |  |
| GRN | 3 (4.1) |  |  |

#### Sensitivity and specificity of the proposed diagnostic criteria

Based on the most common early symptoms in the patients with rATL-predominant degeneration, here we propose a new set of diagnostic criteria for this syndrome (Table 4). To test the sensitivity and specificity of these criteria, we contrasted the rATL- and frontal-predominant patients (Supplementary Table 8), groups that are often difficult to disentangle clinically. To avoid circularity, we did not calculate sensitivity or specificity values based on the neuroimaging data because our groups were anatomically defined.^31,75^ In the first three years of the illness, the criteria differentiated the rATL-predominant from the frontal-predominant group with a sensitivity of 81.3% and a specificity of 84.2%. The sensitivity increased to 86.0% at the time of the first clinical visit and to 93.0% when considering symptoms across all visits. The specificity was 82.8% at the first clinical visit and 81.4% when considering all visits. We predict that sensitivity and specificity will increase in prospectively collected samples because of the increased probing of nonverbal socioemotional semantics during patient evaluations, and should improve further with the inclusion of patients’ neuroimaging information.

**Table 4:**
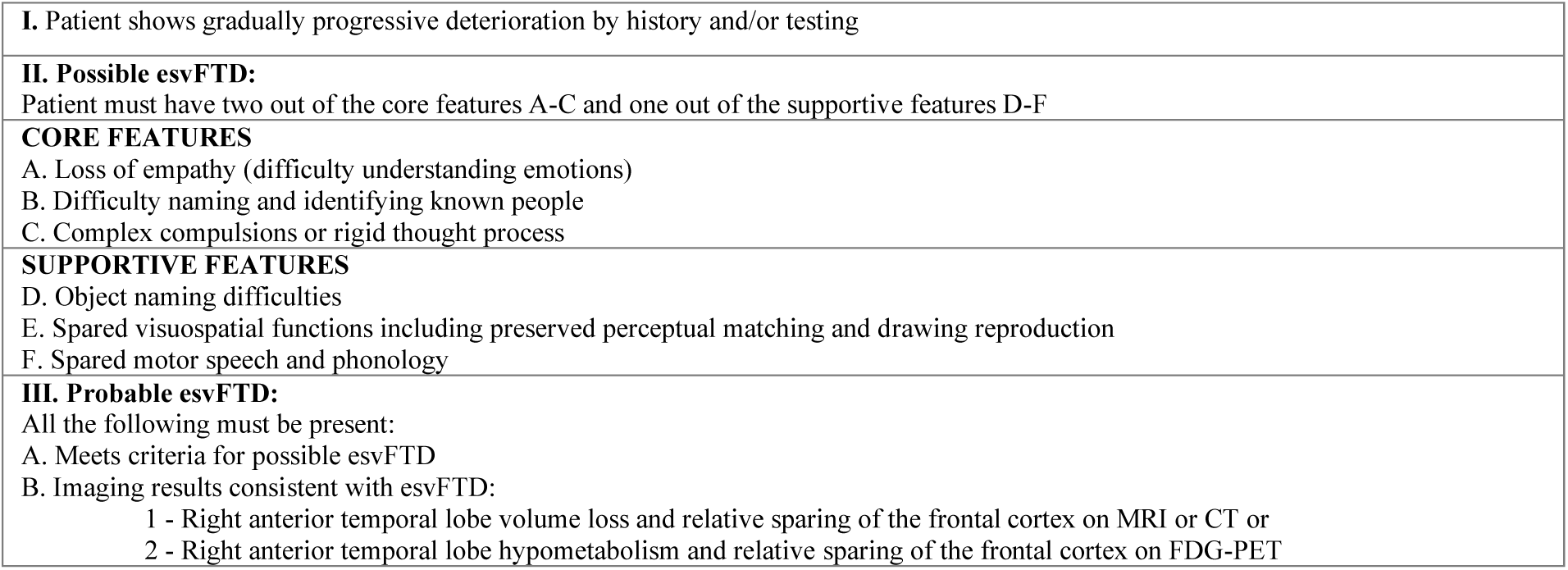
Proposed diagnostic criteria for emotional semantic variant frontotemporal dementia (esvFTD)

The differential diagnosis of rATL-from lATL-predominant patients is somewhat less difficult in clinical practice, in part because all lATL patients show early word-finding and word-comprehension deficits instead of early behavioral symptoms, and thus are easily classified as having a PPA syndrome. As the disease progresses and neurodegeneration spreads to the rATL and orbitofrontal regions,^4,13,14^ however, the continuum between the two clinical presentations becomes more obvious, as predicted by the same FTLD-TDP pathology. Consistent with this, the proposed criteria showed 76.0% sensitivity and 87.0% specificity in distinguishing rATL-from lATL-predominant patients in the first two years of symptoms and 81.3% sensitivity and 68.2% specificity by the third year. The decrease in specificity by the third year highlights the overlap in disease progression between rATL and lATL degeneration.

## Discussion

Here, we present the symptom chronology, neuropsychology, and socioemotional features of a large cohort of well-characterized patients with predominant rATL degeneration. These patients demonstrated early loss of person-specific semantic knowledge (i.e., mainly face-based, nonverbal semantic knowledge) and empathy as well as complex compulsions and rigid thought process, while later they showed loss of verbal semantic knowledge and eventually apathy and disinhibition. Cognitive and anatomical data showed that rATL-predominant degeneration disrupts the neural representations sustaining mainly nonverbal semantic knowledge for socioemotional concepts, resulting in early, prominent deficits in empathy, people recognition, and social behavior. This constellation of symptoms reflects dysfunction in underlying neuroanatomical systems that are anchored to the rATL and that overlap with, but are dissociable from, those involved in frontal-predominant bvFTD and lATL-predominant svPPA.^76,77^ As such, this syndrome necessitates a distinct nomenclature, which herein we refer to as “emotional semantic variant frontotemporal dementia” (esvFTD), a term that reflects the continuum with the semantic variant PPA (and its original semantic dementia), while highlighting socioemotional cognition and behavior, and not aphasia, as the most common first symptom. The neuropathological etiology of this clinico-anatomical syndrome is usually FTLD-TDP disease, and most often FTLD-TDP type C. The intention of using this name is to describe both the behavioral and cognitive symptoms of this syndrome rather than use an anatomical label, such as in “right temporal variant,” which may also be inaccurate in non-right-handed individuals. In these clinical criteria, loss of nonverbal socioemotional semantic knowledge is the central deficit (Table 4). The core features include loss of empathy, difficulty identifying and naming known people, and complex compulsions or rigid thought process. Supportive features include object naming difficulties, spared visuospatial functions, and preserved speech production (motor speech and phonology). A diagnosis of probable esvFTD also requires neuroimaging evidence of disproportionate rATL atrophy or hypometabolism. The novel diagnostic classification proposed here help identify symptoms that are most specifically associated with rATL degeneration. Although the socioemotional evaluations sensitive to FTD can be complicated by the cognitive difficulties,^78^ this study suggests that including certain measures such as Famous Faces, CATS, PPVT, PPT-P, emotional and cognitive Theory of Mind, IAS-Coldness, and TASIT in the assessment of patients presenting with behavioral and language symptoms is useful in diagnosing patients with esvFTD.

As the rATL is a key hub in socioemotional semantic knowledge—the sensorimotor activities, visceral changes, and subjective experiences that are bound into multimodal concepts,^31,45^ we expected that patients with rATL-predominant degeneration would have core deficits in person-specific knowledge. Socioemotional semantic deficits would interfere with their ability to recognize familiar others, attribute meaning to their emotional expressions, and respond appropriately in social contexts.^4,10,11,13,23^ Identification of known people from their face or voice requires person-specific semantic knowledge that incorporates visual and auditory information about what they look and sound like with biographical information about who they are and how they relate to the observer. Consistent with our hypotheses, patients with rATL-predominant degeneration displayed severe impairment on the Famous Faces Naming, Semantic Association, and Familiarity tasks, a pattern consistent with previous reports.^20^ These deficits differed from classical prosopagnosia, the visual inability to recognize familiar people from their faces only, as the patients with rATL-predominant degeneration were also unable to recognize familiar others from their face, name, voice, biography, or information about their relationship to the patient.^28,42^

Loss of empathy was another hallmark feature of patients with rATL-predominant degeneration. Informants who had been close to the patient before disease onset also reported loss of cognitive and emotional empathy, lack of responsiveness to others’ emotional expressions, and increased interpersonal coldness (despite preserved interpersonal dominance, a personality profile that distinguished them from the lATL- and frontal-predominant groups). On socioemotional testing, patients with rATL-predominant degeneration had difficulty selecting a label for facial emotional expressions on the CATS emotion identification task, despite preserved face perception on CATS face identity-matching. This pattern differed from the frontal-predominant group who exhibited impaired performance on both subtasks, and the lATL-predominant group who showed no impairment on both subtasks. Understanding others’ feelings also requires semantic knowledge about nonverbal stimuli (tone of voice, body position, facial expression) as well as access to autonomic, bodily cues that foster vicarious experience of others’ internal states.^79,80^

These findings suggest patients with rATL-predominant degeneration have impaired understanding of the semantics of observed facial expressions and are consistent with literature showing disrupted face processing in both lATL- and rATL-predominant patients.^81^ Patients with rATL-predominant degeneration also had difficulty labeling emotions in videos of socioemotional scenarios in the TASIT-EET. Although the videos included facial, prosodic, postural, and gestural emotional cues, patients with rATL-predominant degeneration performed poorly on this task, suggesting a multimodal nonverbal loss of emotional cue comprehension. Moreover, patients with rATL-predominant degeneration were impaired at interpreting videos that tested emotional Theory of Mind, despite the fact that the emotions of the characters were explicitly verbally labeled for them throughout the task. Of note is that patients had no trouble interpreting cognitive Theory of Mind videos that relied on perspective-taking focused on physical objects rather than on others’ changing emotions. This was in contrast to the frontal-predominant group, who demonstrated impairment on both the cognitive as well as the emotional tasks. Taken together, these findings suggest that rATL-predominant patients’ deficits were due to problems understanding emotions, rather than a result of task-specific non-emotional cognitive demands, and showed that their emotion comprehension deficits extend deeper than the retrieval of the name of the emotion.

rATL- and lATL-predominant atrophy patients commonly exhibited complex, goal-oriented, and time-consuming behaviors and cognitive rigidity. Frontal-predominant patients, however, showed more simple motor repetitive behaviors, hoarding, and obsessions. Compulsive behaviors in bvFTD commonly include aberrant motor behaviors such as tapping and pacing, hoarding, and echolalia and localize to frontal subcortical networks and left lateral temporal lobe.^82,83^ Whereas more complex compulsions such as preoccupation with certain ideas or activities, following fixed schedules, parsimony, and complex rituals were shown to localize to the rATL.^84^ Consistently, our rATL patients most commonly exhibited complex, goal-oriented, and time consuming behaviors and rigid thought process.

In the later phases of disease, patients with rATL- and lATL-predominant degeneration also exhibited apathy and disinhibition, symptoms that are cardinal features of bvFTD. In the frontal-predominant group, conversely, apathy, disinhibition, and poor judgment/dysexecutive symptoms were the most common early symptoms. This suggests that the early symptoms can help distinguish temporal-from frontal-predominant FTD. In FTD, apathy can reflect underlying deficits in cognitive, behavioral, or affective systems that are anchored by the frontal lobes.^55^ Disinhibition in FTD can refer to behaviors that reflect a lack of impulse control, imbalanced reward and punishment systems, executive dysfunction, lack of disgust, or difficulty understanding emotional contexts.^85^ The clinical histories of rATL-predominant patients in our cohort suggested that symptoms recorded as apathy and disinhibition differed from typical examples reported in the frontal-predominant group and commonly known for bvFTD. For instance, lack of participation in activities with family or making tactless comments were due to socioemotional semantic deficits rather than apathy or impaired impulse control. By history, the rATL-predominant degeneration patients had early loss of interest in friends and family, were less affectionate, and made tactless comments indicating a disregard for the social context but did not show deficits in impulse control until after year four of illness.

Early semantic dementia descriptions focused on the prominent verbal semantic deficits resulting primarily from lATL damage,^6,86,87^ the recent use of more comprehensive neuropsychological batteries has yielded data clearly delineating the socioemotional semantic impairments that predominate in the setting of rATL degeneration. The homogeneous pathology in esvFTD patients emphasizes the clinical importance of separating this group from the more pathologically heterogeneous bvFTD, while placing it on a continuum with its svPPA counterpart. esvFTD (behavioral syndrome) and svPPA (language syndrome) are the two extremes of the semantic dementia continuum. Recognizing both the behavioral and linguistic challenges linked to focal ATL atrophy is important to isolate this relatively homogeneous clinico-anatomical-pathological syndrome from the more pathophysiologically heterogeneous frontal presentations. Consistent with studies that showed emotional deficit in svPPA,^16,88^ our findings highlight that socioemotional changes are present in lATL-predominant patients, although they are not the main complaint and are less prominent than in rATL (see Figure 3). Given the common but graded symptomatology between rATL- and lATL-predominant patients, the proposed criteria can be considered an extension of the svPPA definition and refinement of the original semantic dementia description. As disease progresses, the left and right presentations will often merge, and most patients will develop both language and comportment difficulties. The symmetry between lATL and rATL degeneration is likely not a perfect mirror image, however, as one longitudinal study indicated that while lATL patients demonstrated progressive atrophy in the rATL, rATL patients showed further atrophy in right hemisphere regions including orbitofrontal cortex and anterior cingulate cortex.^13^ Stronger functional connectivity between the ATL and orbitofrontal regions in the right hemisphere has been found in healthy individuals, which may help to explain these different effects.^89^ While the challenge of mapping the clinical and pathological continua across all FTD-spectrum disorders is well-known and accepted,^75,90–93^ clear diagnostic algorithms are needed to improve early diagnosis, pathological prediction and specialized care and treatment.

The high frequency of the first two symptoms in the rATL-predominant cohort (loss of empathy and person-specific semantic knowledge) and the deficits on both facial emotion identification and famous faces suggest that the regions subserving face and emotion processing are interlinked and possibly undergo interdependent development during maturation and concordant decay during neurodegeneration. Neurodevelopmentally, the ability to acquire and respond to social and emotional concepts is shown to be linked to accurate interpretation of emotional expressions during early childhood. In fact, recognition of emotional facial expressions is a fundamental aspect of human behavioral neurodevelopment, as infants prefer to look at faces from a very early age, and regulate their actions based on maternal emotional facial expressions.^94^ Furthermore, impairment in recognizing emotional facial expressions is presumed to be one of the mechanisms underpinning the behavioral symptoms in autism spectrum disorder which involves the rATL.^95–98^ Recent work proposes that developmental factors might influence vulnerability to specific neurodegenerative illnesses and links to specific phenotypic presentations.^99,100^ In particular, previous studies suggest that non-right-handedness is over-represented in svPPA compared to other PPA variants and to the general population.^99^ In our esvFTD cohort, there was also a relatively high prevalence of non-right handedness (15%) compared to the 10% reported in the general population.^101^ Furthermore, a previous case report described a behavioral presentation in a non-right-handed patient who had left temporal predominant atrophy.^48^ Taken together, this evidence suggests that handedness, and thus lateralization of language and emotion processing, might influence how linguistic and behavioral symptoms associate with ATL atrophy, contributing to phenotypic variation.^99,100^

Although loss of empathy is the most common symptom used by clinicians and caregivers to describe the early stages of rATL-predominant degeneration, a previous study suggested a prodromal phase of irritability, emotional distance, and changes in sleep, appetite, and libido.^4^ In the present study we considered the subtle early emotional changes such as becoming more selfish and emotionally distant as part of loss of empathy as these symptoms are likely the subtle early manifestations of socioemotional semantic loss. Libido changes and irritability happened in the context of loss of empathy. Similarly, appetite changes happened in the context of other complex compulsions. Sleep changes happened as a prodromal symptom only in a minority of patients. It is possible that the prevalence of these symptoms is underestimated and masked by the more pressing symptoms by the time patients present for evaluation.

A recently proposed diagnostic framework for focal rATL degeneration identified memory symptoms and prosopagnosia as key features but did not distinguish between episodic or semantic memory.^102^ Our results suggest the deficits in the rATL-predominant group extend beyond classical prosopagnosia and represent a multimodal semantic loss for person-specific concepts, but it is also possible that some rATL-predominant patients may have selective prosopagnosia (without person-specific knowledge) in the very early stage of their illness.^103^ We believe that our large sample size, and comprehensive language and socioemotional testing battery enabled us to derive a more complete and precise depiction of symptoms, while at the same time highlighting a semantic memory deficit as the common underlying mechanism in both esvFTD and svPPA. Defining the nature of memory loss in these patients as mainly semantic rather than episodic is particularly relevant, because including episodic memory deficits as a core diagnostic criterion for rATL degeneration syndrome is likely to cause diagnostic confusion with clinical AD, particularly in settings where AD biomarkers are unavailable.

This study has several limitations. Although our cohorts of patients were studied prospectively by a multidisciplinary team that included experts in both language and behavior, the retrospective nature of chart reviews is heavily informed by the clinician writing the original report. Given our reliance on patients’ and caregivers’ recollections of the symptom chronology, it is possible that recall bias may have influenced our findings (though our large sample size makes this less likely). Although it is difficult to ascertain the natural history of early symptoms in rare diseases such as the ones we study here, future collaborative, prospective cohort studies with shared measures and approaches will be imperative for making strides in this area. Another limitation pertains our imaging-based selection criteria, which focused on identifying rATL-predominant cases and excluded cases that had both concomitant severe frontal or lATL atrophy. Additional studies are needed to phenotype the subset of FTD patients with other patterns of atrophy, such as co-occurring right frontal and bilateral temporal damage. In addition, although the rATL syndrome we describe includes prominent atrophy in this area, these patients also have atrophy in a network of regions (e.g., lATL and right insula) connected to the rATL. The atrophy of each group as shown in Figure 2 extends beyond the regions of maximum atrophy and as such symptoms could be due to atrophy in the other regions involved or multiple parts of the connected networks. Future work is needed to further elucidate how structural and functional damage in the rATL and its associated networks relate to the symptoms we detected in this group. Finally, we acknowledge the serious limitation that patients included in this study are mostly white, highly educated, and native English speakers. Further studies that include more diverse patient populations are needed to shed light on the cultural and environmental variability of socioemotional and linguistic presentations in patients with focal ATL degeneration,^104^ and will provide clinical tools necessary to capture culture-specific language and emotional stimuli.

In conclusion, we show that patients with rATL-predominant degeneration show early loss of empathy and person-specific knowledge in relation to damage to the neural systems responsible for non-verbal, socioemotional semantic knowledge. Progression of disease includes language-based semantic loss, highlighting the continuum with the semantic variant of PPA and the original description of semantic dementia. In an effort to improve precise communication, we propose the term “emotional semantic variant frontotemporal dementia (esvFTD)”, emphasizing the cognitive (semantic) and chief clinical (behavioral) features of this disorder. Specific neuropsychological tests that investigate knowledge of emotions, social concepts, and biographical information for known people are important for capturing early esvFTD symptoms and should be included in standard evaluations. Accurate identification of esvFTD patients will pave the way to better prognostication and therapeutics and will help to advance our understanding of the role of nonverbal semantics in human social behavior.

## Data Availability

The data for this study is available upon request.

## Abbreviations

bvFTD: behavioral variant frontotemporal dementia
esvFTD: emotional semantic variant frontotemporal dementia
FTD: frontotemporal dementia
FTLD: frontotemporal lobar degeneration
rATL: right anterior temporal lobe
lATL: left anterior temporal lobe
sd: standard deviation
svPPA: semantic variant primary progressive aphasia
TDP-43: transactive response DNA binding protein 43

## Acknowledgements

The authors thank the research patients and their families for the time and effort they dedicated to research at UCSF’s Memory and Aging Center. VBM analyses were performed using the Brainsight system, developed at UCSF by Katherine P. Rankin, Cosmo Mielke, and Paul Sukhanov, and powered by the VLSM script written by Stephen M. Wilson, with funding from the Rainwater Charitable Foundation and the UCSF Chancellors Fund for Precision Medicine. We thank Anna Karydas and Jennifer Yokoyama (UCSF Memory and Aging Center genetics experts) for assistance with genetic data, as well as Dr. John Trojanowski (Department of Pathology and Laboratory Medicine, University of Pennsylvania. Pennsylvania, Philadelphia) for assistance with autopsy data.

